# Non-Invasive Multiplexed Real-Time Threshold Detection of Cardiac, Renal, and Metabolic Biomarkers via Wrist-Worn Infrared Spectroscopy

**DOI:** 10.64898/2026.01.15.26344213

**Authors:** Jitto Titus, Jessie Katz, Karina Soto-Ruiz, Robert Christenson, Alan HB Wu, Allan S. Jaffe, W. Frank Peacock

**Affiliations:** RCE Technologies; Comprehensive Research Associates; University of Maryland School of Medicine; Zuckerberg San Francisco General Hospital; Department of Laboratory Medicine and Pathology at Mayo Clinic; Baylor College of Medicine

**Keywords:** Transdermal sensor, Troponin I, BNP, HbA1c, Creatinine

## Abstract

**Object:** Transdermal biosensors may provide an alternative to conventional blood-based biomarker measurement. Our purpose was to determine the binary correlation between transdermal (Infrasensor™; RCE, Inc, Carlsbad, CA) and conventional blood-based measurements.

**Methods:** This was a secondary analysis from a previously published observational cardiac troponin I (cTnI) study performed to establish the upper reference level of cTnI, at 10 US hospitals. After obtaining informed consent, 2 cohorts of patients were enrolled: 1) those who completed a health assessment questionnaire and appeared healthy, and 2) those with a known elevated cTnI per the local hospital standard assay. All blood lab analyses were performed at the University of Maryland Medical Center, Baltimore, MD. Normal was defined as cTnI <53.48 ng/L (male) or 34.11 ng/L (female) using the Siemens Atellica IM assay (Siemens Medical Solutions, Mountain View, CA), NT-proBNP <450 pg/mL (>75 years) or <124 pg/mL (<75 years), creatinine >1.17 mg/dL (male) or >0.95 mg/dL (female), and HbA1c <6.4%. The Infrasensor was placed on the patient’s wrist for measurement and blood drawn for analysis at approximately the same time.

**Results:** Of 840 enrolled patients, the median (IQR) age was 46 (30,57), 416 (49.5%) were female, 10.36% Hispanic, 6.7% Asian, 12.9% African American, and 69.1% White. Elevated lab tests were 102 hscTnI’s, 156 NTproBNP’s, 37 HbA1C’s, and 163 creatinine’s. Significant binary correlations were found between all transdermal signals and the corresponding lab blood levels

**Conclusion:** Infrasensor transcutaneous measurement demonstrates similar results as that obtained from blood testing in the central laboratory.

**Capsule:** The Infrasensor (RCE, Inc, Carlsbad, CA, USA) is rapid point of care transcutaneous biomarker measurement device. This study evaluated its ability to provide qualitative results for troponin I, NTproBNP, creatinine, and HbA1c levels in 840 patients. Significant correlations were found between all transdermal signals and the corresponding binary lab blood levels.

## Introduction

Cardiovascular disease is a leading cause of global morbidity and mortality worldwide (1). Early detection using blood-based monitoring cardiac biomarkers, e.g., high sensitivity cardiac Troponin I (HscTnI) and N-terminal Pro B type Natriuretic Peptide (NT-proBNP), are important adjuncts for diagnosis and management of cardiovascular disease. (2) Recent advancements in technology now allow for the use of novel methods for monitoring physiological parameters, potentially including bloodless biomarkers. (3, 4) Transdermal sensors offer a non-invasive approach to acquire these data at the point of care, as well as continuous monitoring, irrespective of the clinical situation. (5) This study investigates the relationship between transdermally derived signals and blood-based biochemical markers, aiming to enhance assessment strategies, and potentially complement existing blood-based biochemical assays which are presently the state-of-the-art methods for clinical care.

The intersection of transdermal biomarker signals and blood-based biochemical measurements presents a potentially important area of research with significant implications for healthcare and biomedicine. While generic transdermal sensors primarily capture data related to skin properties and local physiological changes, the ability to leverage this approach to detect blood-based markers offers a potentially comprehensive view of systemic biochemical dynamics. (2, 5) Recent innovations in wearable sensor technology can leverage the capabilities of infrared optics fashioned to perform molecular spectroscopy and may provide biomarker signatures more rapidly. Exploring the associations between transdermally derived signals and blood-based measurements holds promise for enhancing diagnostic accuracy and speed, patient satisfaction, as well as providing prognostic capabilities in personalized medicine. (4, 6).

A new device, called an Infrasensor™ (RCE, Inc, Carlsbad, CA), reports near instantaneous signals related to biomarker levels with a wrist-worn transdermal infrared spectrophotometric sensor that does not require blood sample collection. (7) See figure 1. The infrared sensor ranges for the biomarkers of interest were determined by interrogation of purified material for each of the analytes involved. (7,8) The Infrasensor™ is comprised of 4 detectors or channels sensitive to 4 different wavelength ranges. Troponin optical signatures are found in all ranges, however by design, one of the channels has a larger contribution from troponin than the other analytes. Similarly, there is optical information corresponding to NT-proBNP, Creatinine, and Glycated Hemoglobin (HbA1C) present across the 4 wavelength ranges with varying degrees of contribution. The 4 channels of the signal can be combined with each having a unique weight multiplier as informed by the Machine Learning (ML) model’s ability to best predict the “ground truth”. Hence distinct ML models can be created for different biomarkers. See figure 2. Translational research and early biochemical validation efforts show reasonable binary correlations with HscTnI (9, 10), and potential associations with phenotypes of acute cardiovascular disease of clinical relevance. (11, 12) Other biomarkers have not previously been explored prior to this investigation.

**Figure 1.**
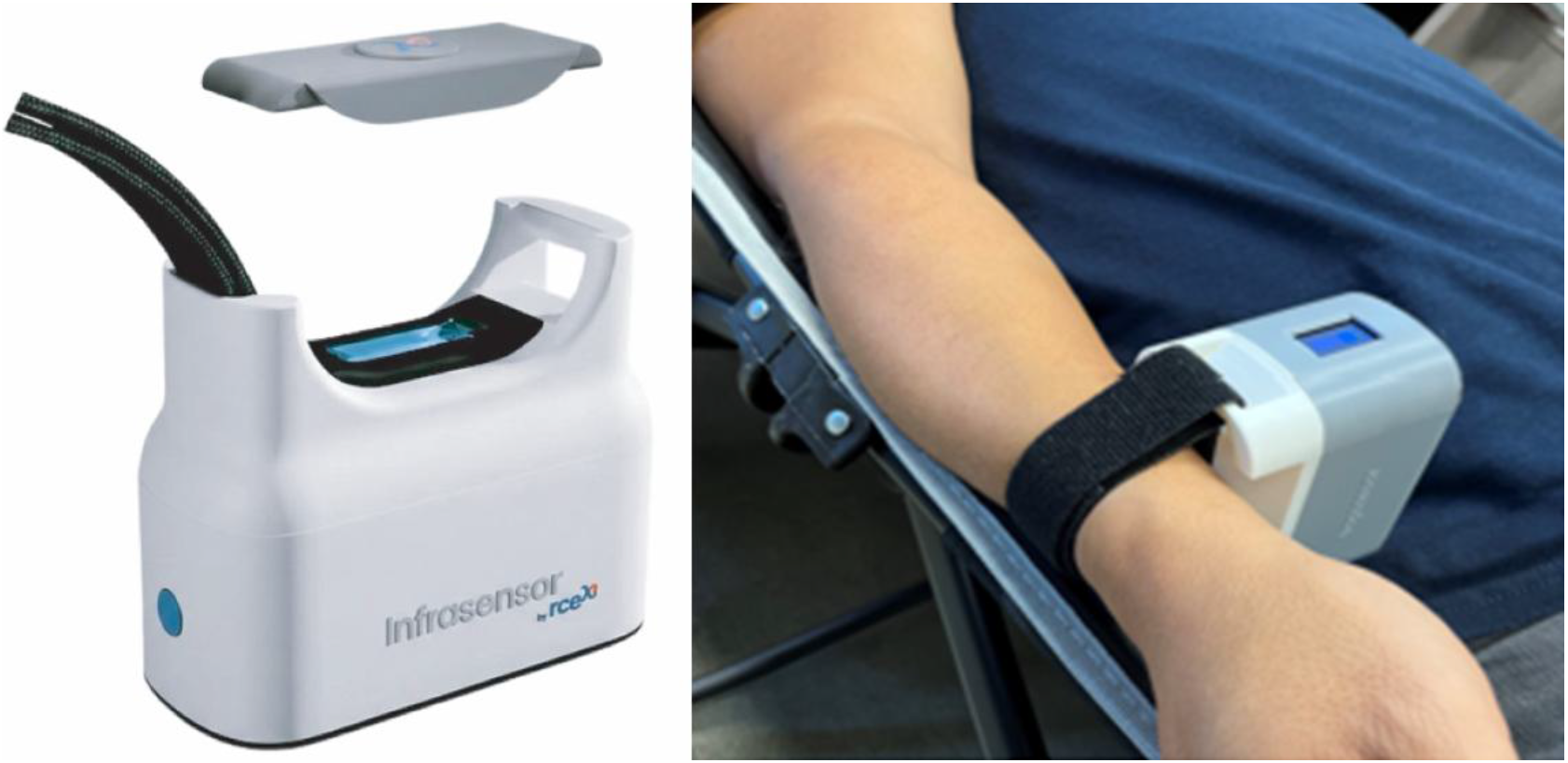
Infrasensor

**Figure 2.**
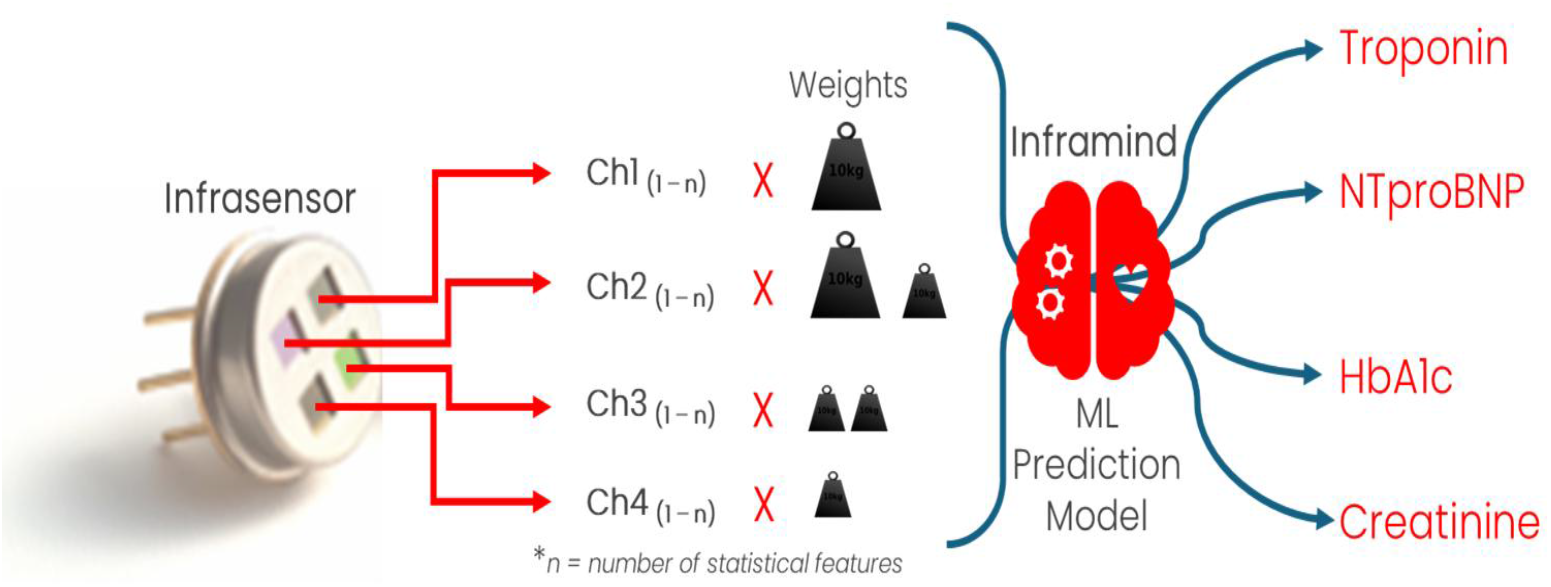
Infrasensor Waveform coupled with feature weights for biomarker association using a machine learning model

In an effort to further understand the associative capabilities of the Infrasensor™, using biomarkers of cardiac and metabolic origin, a multisite study enrolling healthy individuals as the control group, and a comparator group with known myocardial injury, was conducted.

## Methods

We performed a prospective observational cohort evaluation of the Infrasensor™ to determine binary correlation with common local lab measurements. Conducted at 10 hospitals across the US, each with ethics committee approval, patients were enrolled into 2 cohorts. A healthy control cohort was expected to have normal troponin results, and was formed to establish the baseline range for the Infrasensor™. Enrollment criteria mandated that participants complete a questionnaire regarding health conditions. Exclusion criteria included self-reported cardiac conditions, hospitalization in the past 3 months, thyroid disease, current pregnancy, and ongoing treatment for diseases including cancer, liver, lung, or autoimmune disease. The second cohort consisted of hospitalized patients with confirmed elevated local laboratory cTnI as part of their routine clinical care, defined as a TnI concentration exceeding the local institutions standard of care 99th percentile upper reference limit (URL). The overall sample size was prespecified to be at least 800 patients, as consistent with URL definition recommendations. (13)

If inclusion criteria were met, and after written informed consent obtained, Infrasensor™ measurements and blood draws were obtained as close together as possible. Blood samples, processed within one hour, were stored at −80°C until being shipped to the CCRL (Clinical Core Research Laboratory) Facility at the University of Maryland School of Medicine, Baltimore, MD, a CLIA (Clinical Laboratory Improvements Amendments) licensed and CAP (College of American Pathologists) accredited laboratory, for measurements of Hs cTnI I (Atellica IM TnIH assay, Siemens Healthineers, Terrytown, NY)), NTproBNP, HbA1c, and creatinine. Normal results were defined as HscTnI <53.48 ng/L (male) or 34.11 ng/L (female), NT-proBNP <450 pg/mL (>75 years) or <124 pg/mL (<75 years), creatinine <1.17 mg/dL (male) or <0.95 mg/dL (female), and HbA1c <6.4%.

Statistical analyses, including logistic regression and decision tree-based modeling, were utilized to create a binary classification between transdermal and blood-based markers. A gradient-boosting tree-based model was applied to address the binary classification problem of discriminating between elevated and non-elevated biomarkers. The model leveraged the features extracted by RCE’s Infrasensor™, which provided a dimensionality reduction that proved advantageous for tree-based machine learning algorithms. This reduced feature space led to enhanced performance compared to more complex network-based models, which often struggle with high-dimensional data due to the ‘curse of dimensionality’. (14) Gradient Boosting is an ensemble machine learning technique that iteratively combines weak learners, typically decision trees, to create a robust predictive model. The algorithm works by sequentially fitting decision trees to the residuals of the previous trees, allowing subsequent trees to focus on the samples that were misclassified by the preceding models. To implement the Gradient Boosting model, the XGBoost library was utilized. (15)

The evaluation of Infrasensor™ performance followed a five-fold cross-validation method which is a statistical technique used to evaluate the performance and generalizability of a machine learning model. With this strategy the dataset is randomly divided into five equal-sized subsets or “folds”. The model is trained and validated five times, each time using a different fold as the validation set and the remaining four folds as the training set. This process ensures that every data point is used for both training and validation, providing a more accurate assessment of the model’s performance. The healthy and elevated samples were allocated equally across the five folds. Employing this approach minimizes variability arising from random train-test split selection, thereby enhancing the reliability of assessing the model’s performance. The results were reported with 95% confidence intervals (CI) and C statistics. The performance of the AI-trained model (from the Infrasensor™ data) was correlated with plasma-based core lab results, using predefined cutoff points, and was evaluated for sensitivity, specificity, positive predictive value, and negative predictive value.

## Results

Overall, 840 patients were enrolled. Of these, 416 (52.4%) were female,10.36% Hispanic, 6.7% Asian, 12.9% African American, and were 69.1% Caucasian. Elevated levels of markers were found in 102 (12.1%) hscTnI, 156 (18.6%) NTproBNP, 37 (4.4%) Hba1C, and 163 (19.4%) creatinine in the patients for whom the tests were done. See table 1.

Our analysis revealed significant binary correlations between transdermally derived signals and blood-based levels of HscTnI, NT-proBNP, HbA1c, and creatinine, see figure 3. Promising results were found regarding the performance of the Infrasensor™ in predicting elevated troponin levels (AUC = 0.95), particularly when sex was considered. A significant binary correlation was identified between transdermal signals and NT-proBNP (AUC = 0.91). Additionally, a positive binary correlation was found between transdermally derived signals and HbA1c levels (AUC = 0.84) and those with creatinine (AUC = 0.87).

**Figure 3.**
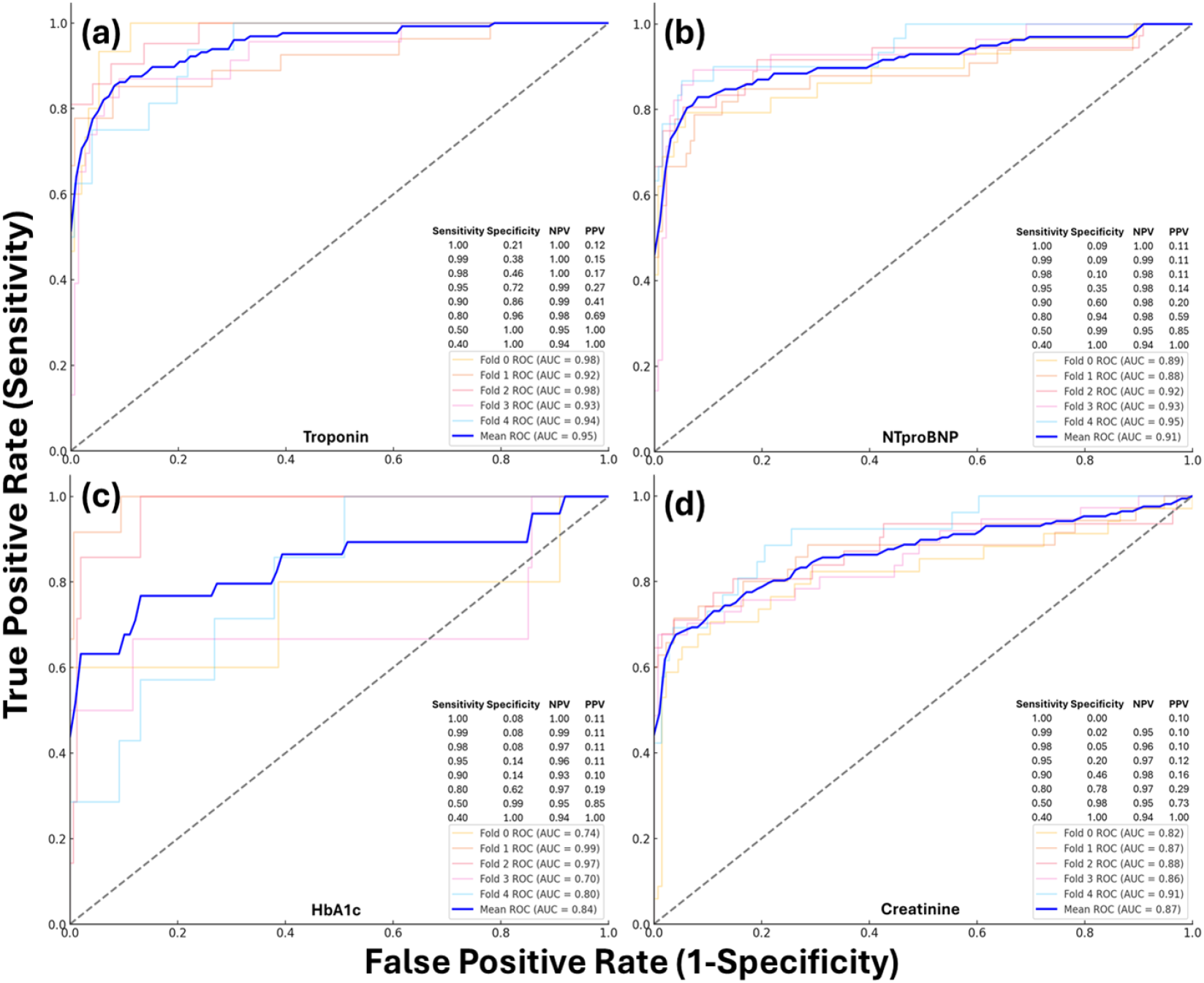
Biochemical Validation with hs-cTnI, NTproBNP, HbA1c, Creatinine

Because models’ performances are related to biomarkers being concomitant and inter-correlated, due to the presenting pathology, we performed a sensitivity analysis by the creation of a subset of samples where the 4 biomarkers were “exclusively” elevated. See figure 4. Considering the hypothesis that the models are unique and the Infrasensor’s™ 4 channels can be uniquely combined, the model trained to predict a certain biomarker will perform well in predicting that biomarker, but the performance reduces markedly with the others for which it is not directed.

**Figure 4.**
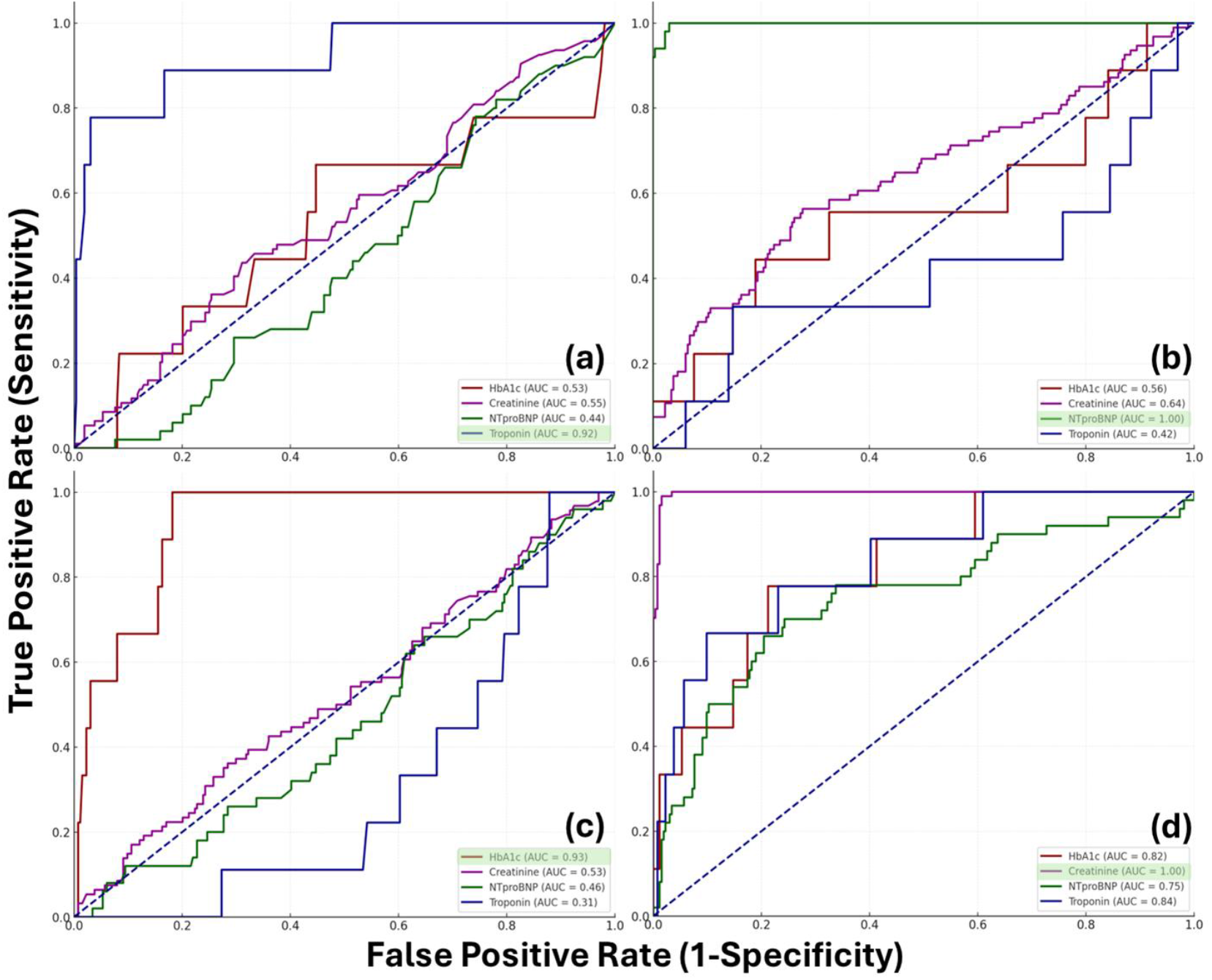
Models trained to predict one biomarker (in green box) and tested for the other biomarkers

## Discussion

Our findings elucidate the potential utility of transdermal monitoring and its association with biomarker levels that can be leveraged for precision medicine and disease phenotyping. The significant binary correlations, particularly with Troponin I and NT-proBNP, suggest that transdermal signals may serve as valuable indicators or complementary measures for a broad range of health conditions. The ability to exclude an increased biomarker for all of the blood analytes is highly effective. However, the specificity and positive predictive values are presently less than ideal, suggesting some other infrared signals are also being captured.

The observed associations between transdermal signals and blood-based biomarkers suggest a promising avenue for non-invasive diagnostics across various medical domains. Transdermal wearable sensors offer real-time monitoring capabilities, potentially enabling early detection or exclusion of health abnormalities. (16) Integration of transdermal data with traditional biomarker assessments could enhance risk stratification and personalized treatment approaches.

One of the notable strengths of transdermal sensors is their non-invasive nature, which can lead to improved patient compliance and comfort compared to invasive blood draws. This aspect is particularly relevant for continuous monitoring, where transdermal sensors can provide ongoing data without the need for repeated invasive procedures. (17) The study’s findings support the notion that transdermal sensors could play a crucial role in personalized healthcare by accelerating access to clinical care and necessary interventions based on real-time biomarker assessment at the point of care.

These data contribute to the growing body of evidence supporting the potential of transdermal sensors in broader health assessment. For example, Biobeat, which has sensors that continuously monitor 13 physiological parameters including HR, blood pressure, cardiac output, and single-lead ECG, has been assessed in in-hospital patients for providing real-time monitoring, triggering alerts, and predicting patient deterioration. (16) Zio AT is an adhesive-patch ECG monitoring device that continuously monitors ECG changes and alert patients when they have preset events based on the AI algorithm; this device has been shown to detect significantly more events than a conventional Holter device in ambulatory situations. (18) Multiple companies have products used for noninvasive continuous blood glucose monitoring. (19) The binary correlations between transdermal signals and blood-based biomarkers highlight the potential for advancing non-invasive monitoring technologies in diverse health conditions. Continued research and development in this area are crucial for realizing the full clinical potential of transdermal sensors and improving patient outcomes.

Our study is not without limitations, which include the need for further validation in larger cohorts and consideration of environmental factors affecting transdermal signal accuracy. While the binary correlations between transdermal signals and blood-based biomarkers are statistically significant, further validation in larger and more diverse cohorts is necessary to establish the robustness and generalizability of these findings. In addition, more granular data regarding the degree of increase or position within the normal reference range relative to these blood biomarkers is needed so that thresholds for use can be identified prospectively. Environmental factors such as temperature, humidity, skin condition, and color may also influence transdermal signal accuracy, highlighting the need for standardized protocols and calibration procedures. (6)

Future research directions should focus on refining the transdermal sensor technology to improve accuracy, sensitivity, and specificity in potential clinical areas of interest. Longitudinal studies, encompassing diverse populations, are needed to provide insights into the stability of transdermal signal patterns over time, considering variability for both the methods and biological variation, which has not been assessed continuously. This would be essential to allow for the monitoring of disease progression or treatment response. Additionally, investigating the clinical applications of transdermal sensors could broaden their utility and enhance their impact on precision medicine and disease phenotyping. (20)

## Conclusion

We found a significant binary correlation between transdermally derived biomarkers and blood-based biochemical measurements performed in a CLIA licensed laboratory, highlighting the potential of transdermal wearable sensors as versatile tools for non-invasive health monitoring. The findings suggest that these sensors could play an important role in precision medicine and disease phenotyping across various medical domains. Future research should focus on refining these technologies, validating findings in diverse populations, and exploring a wide range of clinical applications to enhance patient outcomes and advance personalized healthcare.

## Data Availability

All data produced in the present study are available upon reasonable request to the authors

## Conflict of Interests

WFP: Research Grants: Abbott, Brainbox, Quidel, Roche, Siemens. Consultant: Abbott, Astra-Zeneca, Biocogniv, Brainbox, Bristol Meyers Squibb, Instrument Labs, Janssen, Osler, Quidel, Roche, Siemens, Spinchip, CSL-Vifor. Stock/Ownership Interests: AseptiScope Inc, Brainbox Inc, Biocogniv, Inc, Braincheck Inc, Coagulo Inc, Comprehensive Research Associates LLC, Comprehensive Research Management Inc, Emergencies in Medicine LLC, Prevencio Inc, RCE Technologies, ROMTech.

JT: RCE Employee JK: None

KSR: Comprehensive Research Associates, LLC, employee.

RC: Consultant for Siemens Healthineers, Beckman Coulter, Becton Dickinson, Roche Diagnostics, Sphingotech, Polymedco, Werfen

AHBW: Contracts Abbott, Diazyme, Furimori Advisory: Werfer, BrainBox., RCE, Quidel, Truvian Health, and Babson.

ASJ: Has presently or in the past has consulted for most of the major diagnostic companies, this includes Moderna, Siemens Abbott, Roche, Radiometer, ET Healthcare, Sphingotec, Spinchip, LuminaRx, and he has stock in RCE.

